# Serum SARS-CoV-2 nucleocapsid antigen detection is essential for primary diagnostics of SARS-CoV-2-associated pneumonia

**DOI:** 10.1101/2020.09.24.20200303

**Authors:** Yuri S. Lebedin, Olga V. Lyang, Anaida G. Galstyan, Anna V. Panteleeva, Vsevolod V. Belousov, Denis V. Rebrikov

## Abstract

The article highlights the diagnostic value of SARS-CoV-2 seroconversion in patients with pneumonia based on the results of a retrospective study conducted at the height of the COVID-19 pandemic in Moscow, Russia.

## Introduction

Immunochemical detection of antigens in the blood is widely used in the diagnostics of infectious diseases. The antigenemia tests are usually targeted at blood-borne pathogens (notably chronic viral infections including CMV, HBV, HCV and HIV [1-3]). In respiratory diseases, the penetration of the pathogen components (nucleic acids or proteins) into non-respiratory body fluids is likely, but their detection is usually considered of minor value because of the focal nature of the pathogenesis and its strong association with mucous immunity. Despite that, certain respiratory infections can be efficiently diagnosed and monitored by detection of corresponding antigens in the blood; the examples include galactomannan in pulmonary aspergillosis [4]) and *L. pneumophila* antigen in the urine of Legionnaires’ disease patients [5]). In addition, the use of serological immunoassays for the management of virus-associated nosocomial pneumonias in intensive care unit patients has been reported [4,6].

In this report, we evaluate the SARS-CoV-2 nucleocapsid antigen (N-Ag) and respective antibodies as diagnostic markers and demonstrate the total prevalence of N-Ag seroconversion in SARS-CoV-2-associated pneumonia patients.

## Patients and Methods

The retrospective study was carried out at the Federal Center of Brain Research and Neurotechnologies during the spring of 2020. At the height of the COVID-19 pandemic, the in-patient care facilities of the Center were redesigned for the emergency hospitalization and management of the SARS-CoV-2-associated pneumonia patients. The study involved patients with clinical signs of COVID-19 pneumonia (n = 475) flown by ambulance services from different districts of Moscow. The key criterion for the emergency admission was fever with a combination of two or more clinical signs including body temperature of ≥ 38.5 °C, respiratory rate of ≥ 30 breaths per minute and/or SpO_2_ of ≤ 93% [7]. Serum samples for the study were collected from each patient at least twice (at admission and at discharge) concomitantly with the mandatory blood collection procedure for routine blood tests. A subcohort of the patients (n = 20) volunteered for repeated sampling of the blood for SARS-CoV-2 serodiagnostics on day 3-5 after the admission. The in-patient care lasted 20±2 days. Criteria for the discharge included a reduction in C-reactive protein levels at WBC counts within the normal range (above 3 × 10^9^ L) and, notably, a clear tendency towards regression of characteristic signs revealed by computer tomography: the absence of new ground glass opacities, a decrease in the severity of the corresponding changes in the lung tissue and/or a decrease in the volume or degree of consolidation of the ground glass opacities (no more than three, each within 3 cm along the maximal dimension) [8]. The qualitative RT-PCR detection of SARS-CoV-2 RNA in nasopharyngeal swabs was carried out at least twice (at the admission and at the discharge) with the use of the The SARS-CoV-2/SARS-CoV Multiplex REAL-TIME PCR Detection Kit RZN No. 2020/9948 (DNA-Technology LLC, Moscow, Russia; diagnostic sensitivity 10 copies per reaction, diagnostic specificity 99.8%) according to the manufacturer’s protocol.

### SARS-Cov-2 nucleocapsid antigen (N-Ag)

coding sequence (NCBI accession number 045512.2) was cloned by NdeI/XhoI into pET-30b(+) vector, Novagen (EMD Millipore); the plasmid was expanded in *E. coli* Top10 and transformed into *E. coli* Rosetta (DE3) for N-Ag expression.

The conditions were optimized for 37 °C bioreactor fermentation in 3000 mL of rich media (yeast extract, bacto peptone, glucose and trace salts). After 12 h incubation, the culture was induced with 2.5 mM imidazole for 4 h. The obtained biomass was resuspended in phosphate buffered saline (PBS) in a 1:3 (w/v) proportion and disrupted using APV-2000 homogenizer (Spx Flow, USA) at 1200 bar. The solution was clarified by centrifugation at 12,000 g. The insoluble fraction (inclusion bodies) containing the target protein was washed sequentially with PBS, 2% Triton in PBS and a fresh portion of PBS to remove the residual detergent. The protein was dissolved in 8 M urea with 250 mM NaCl and 50 mM phosphate, pH 10.0. The solution was incubated at +4 °C overnight and centrifuged at 15,000 g; the collected supernatant was filtered and supplemented with 20mM imidazole to prevent the non-specific binding of impurities. The polyhistidine-tagged N-Ag protein was purified by immobilized metal affinity chromatography using a nickel column (High Density Nickel #6BCL-QHNi, ABT, Spain) equilibrated with the urea buffer. The protein was eluted with 250 mM imidazole buffer and filter-sterilized.

### Serum IgG antibodies

against nucleocapsid antigen (N-IgG) were detected by solid-phase enzyme immunoassay. Briefly, the recombinant full length SARS-CoV-2 nucleocapsid antigen produced in *E. coli* (XEMA, Moscow, Russia) was coated onto the surface of polystyrene microwells. The sera pre-diluted 100-fold in the ELISA buffer (0.1% Tween-20 and 1% hydrolyzed casein in 0.1 M PBS) were placed in the microwells for 30 minutes at 37 °C. After 3 washes with 0.1% Tween-20 in 0.9% sodium chloride, the wells were exposed to the conjugate of murine monoclonal antibodies XG78 against human IgG (gamma chain) with horseradish peroxidase (XEMA) for 30 minutes. After 5 washes with 0.1% Tween-20 in 0.9% sodium chloride, the bound enzyme was revealed by addition of the substrate-chromogenic mix (TMB substrate, XEMA). The color development was stopped by 5% sulfuric acid and the optical density (OD) at 450 nm was measured in a plate reader (Multiskan MC, Thermo Labsystems). The internal controls (stabilized human serum containing specific IgG antibodies) were included in all microplates to calculate the positivity threshold OD for each run individually. The results are expressed as positivity indexes (calculated as OD for a sample of interest related to OD for the internal control).

### Serum IgM antibodies

against nucleocapsid antigen (N-IgM) were detected by reverse solid phase enzyme immunoassay. Briefly, murine monoclonal antibody against the mu chain of human IgM (clone X616, XEMA) was adsorbed on the surface of polystyrene microwells. The dilutions of sera (prepared in the same way as for the IgG assay) were placed in the microwells for 30 minutes at 37 °C. After 3 washes with 0.1% Tween-20 in 0.9% sodium chloride, a working dilution of conjugate of recombinant full length SARS-CoV-2 nucleocapsid antigen with horseradish peroxidase in ELISA buffer was added to the microwells for 30 minutes at 37 °C. After 5 washes with 0.1% Tween-20 in 0.9% sodium chloride, the bound labeled antigen was detected by TMB substrate and OD reading. The calculation of positivity index (similarly with the IgG assay) was performed by using the internal IgM+ control.

### Serum N-antigen (N-Ag)

determination was performed by the two-site solid-phase sandwich method using monoclonal antibodies (mAbs) generously gifted by Hytest Ltd, Turku, Finland.

The microwells were coated with the capture mAb (clone NP1510, HyTest). Serum samples or assay calibrators (solutions of nucleocapsid antigen in donor serum in the range 20-2000 pg/ml) were incubated with working dilution of HRP-labeled tracer monoclonal antibody (clone NP1517, HyTest) in ELISA buffer with addition of 10 ug/ml of heterophilic immunoglobulin elimination reagent (HIER-E-010, Fapon Biotech, China) for 2 hours at 37C under continuous 600 rpm shaking in a PST-60HL-4 shaking incubator (Biosan, Latvia). The detection was performed with the use of TMB substrate and OD reading. The calculation of N-Ag concentration was done by calibration curve using the DataGraph data reduction software (Visual Data Tools, Inc.). N-Ag concentrations exceeding upper limit of calibration curve (2000 pg/ml) were shown as 2222 pg/ml in calculations and graphic presentation. Samples with OD readings below the 20 pg/ml, lacking resolution from the zero calibrator, are conventionally assigned 15 pg/ml values in the presented data.

### The reference ranges

were determined by analysis of serum samples collected from healthy donors (n = 250) before Dec 2019. In both assays, the threshold (cutoff) value was set to achieve the 100% specificity (none of the donors being considered positive).

### The study protocol was reviewed and approved

by the Local Ethics Committee of the Pirogov Russian State Medical University (meeting #194 from March 16 2020, Protocol No.2020/07); the study was conducted in accordance with the Declaration of Helsinki.

## Results and Discussion

The immunoassay-based detection of serum N-Ag in combination with its respective antibodies confirmed COVID-19 in 280 patients (59%) of the studied cohort. RT-PCR analysis of nasopharyngeal swabs confirmed COVID-19 in 68% of the patients; the interception constitutes 56% which gives the concordance of 79%.

Several patients (n = 21, 4%) were identified as SARS-CoV-2 negative by both RT-PCR tests and serology, i.e. most likely represented cases on non-SARS-CoV-2-associated pneumonias. The small number of such patients is explained by the fact that the study was carried out at the peak of the pandemic.

According to the data, N-Ag antigenemia is characteristic of the vast majority of the emergency patients with COVID-19 (56%) at the time of admission.

The disease phase-related dynamics of serum N-Ag levels are shown in Figure 1. On discharge, the majority of patients (n = 104, 90%) in the studied cohort showed serum N-Ag levels below the lower limit of calibration curve (< 20 pg/ml). However, several patients shown clearly positive serum N-Ag even on discharge.

**Fig. 1.**
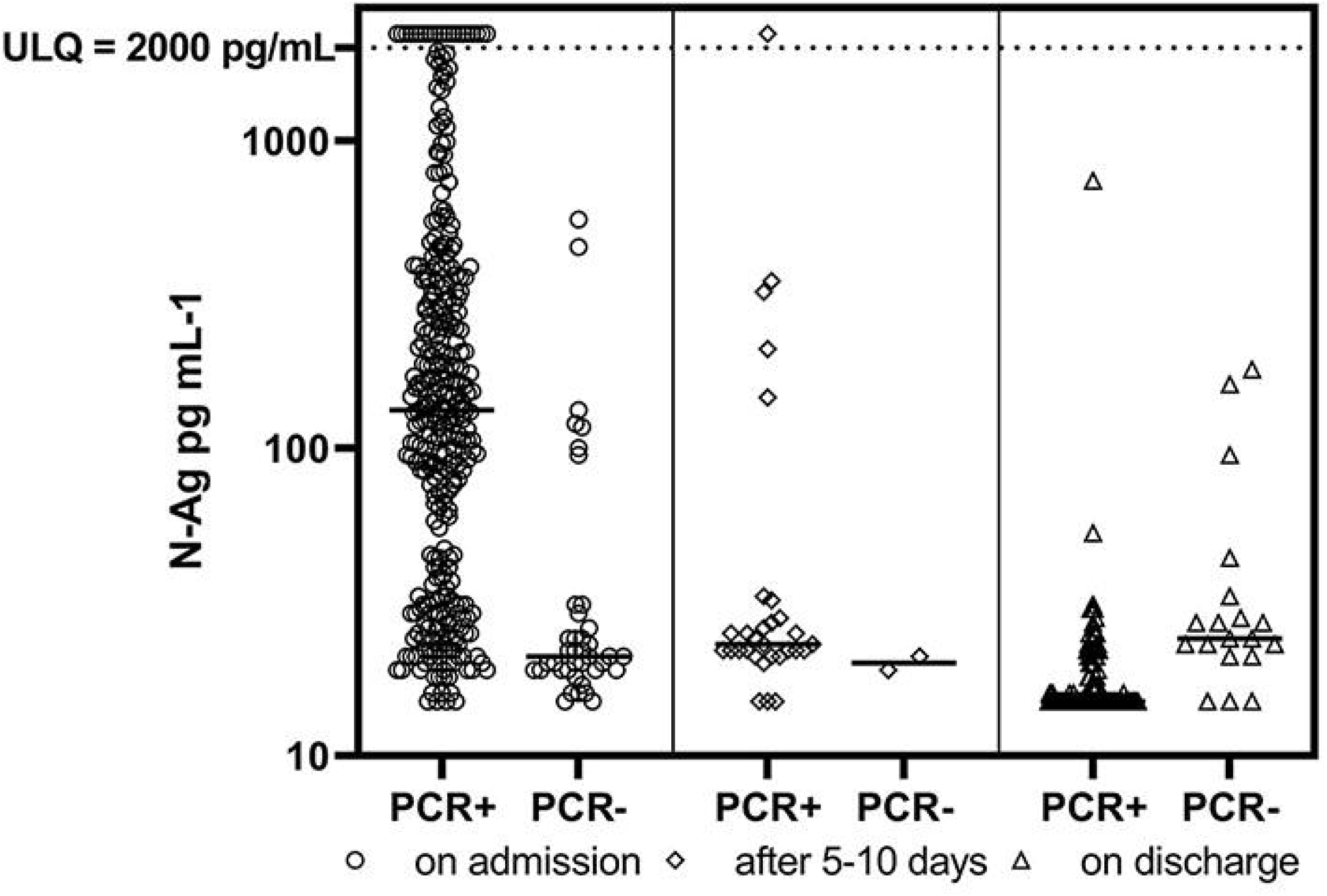
Serum N-Ag levels by disease phase.

By the time of discharge, seroconversion was observed in most of the patients who showed N-Ag positivity on admission (n = 108, 93%), although the degree of seroconversion varied considerably. To monitor the seroconversion patterns, we have compared serum N-Ag with two isotypes of antibody response in serum in individual cases, dividing them in two subgroups by seropositivity on admission (Figures 2ab vs 3ab). The results indicate reciprocal patterns for N-Ag and respective antibodies, which is characteristic of classical seroconversion. The patients showing no serum antibody on admission (presumably being on earlier disease stage) shown more clear pattern of classical seroconversion. The IgG-antibody pattern was more pronounced in both groups than IgM-antibody pattern.

**Fig. 2a.**
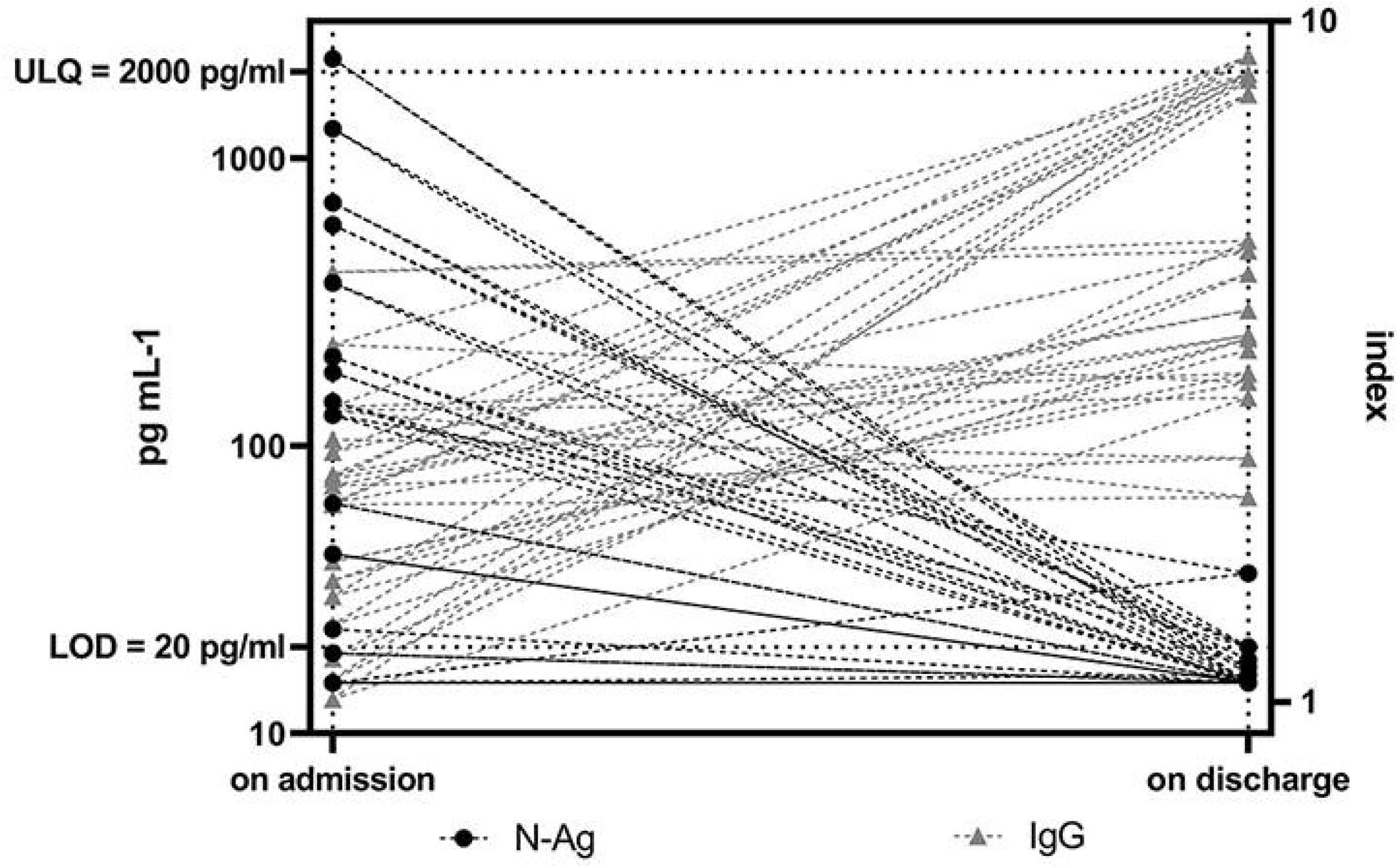
Kinetics of N-Ag and lgG antibodies against N-Ag in patients seropositive on admission.

**Fig. 2b.**
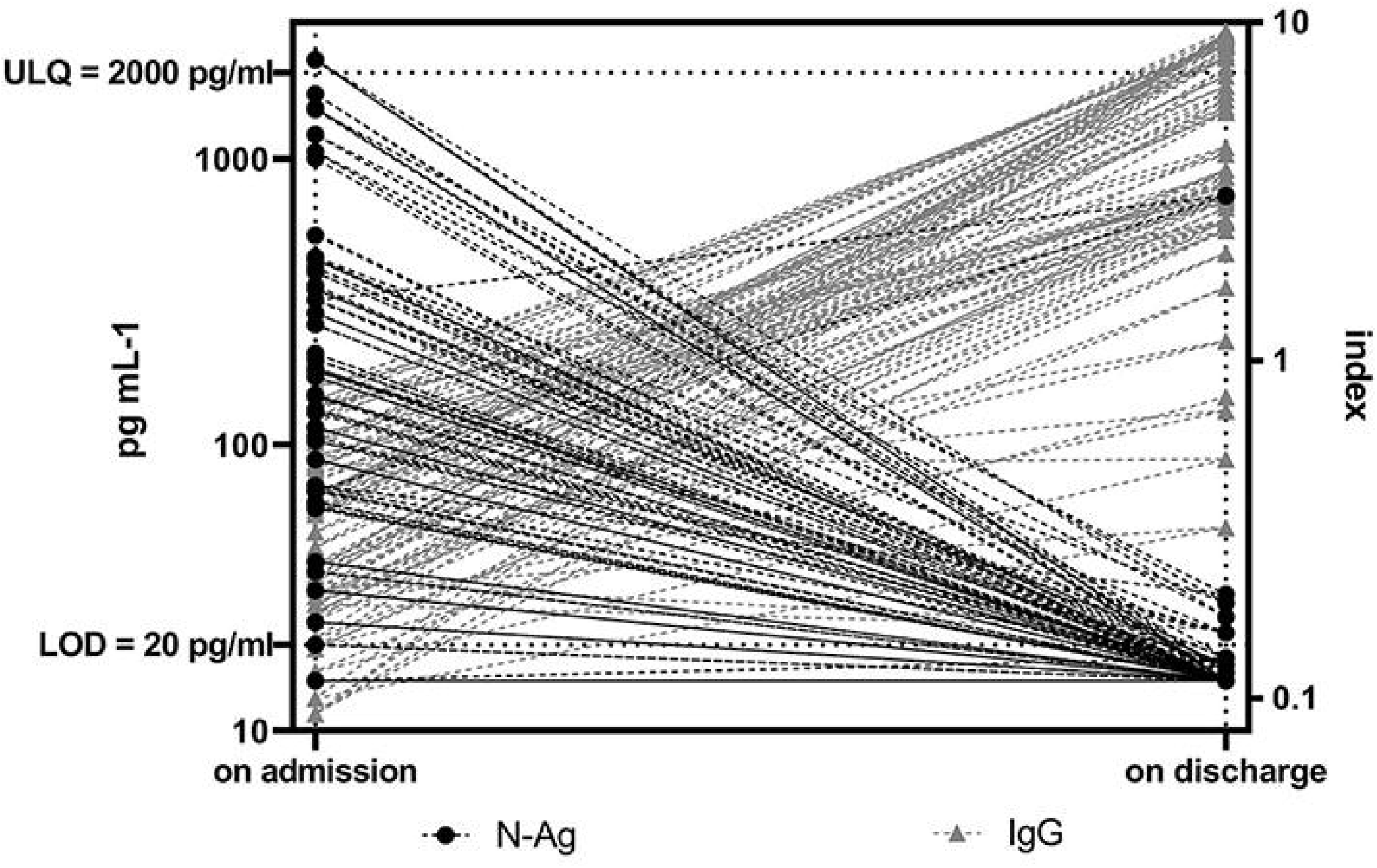
Kinetics of N-Ag and lgM antibodies against N-Ag in patients seronegative on admission.

**Fig. 3a.**
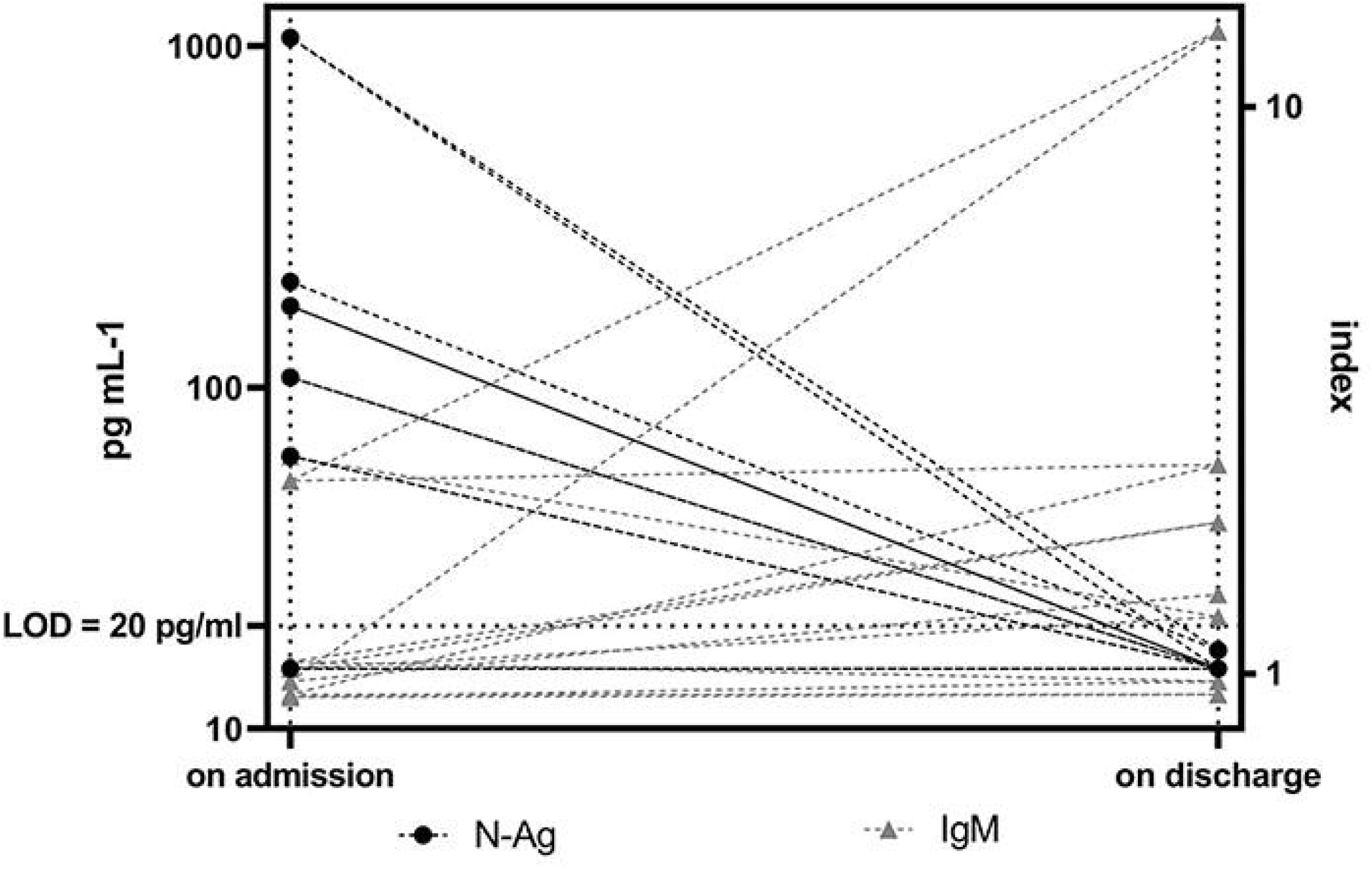
Kinetics of N-Ag and lgM antibodies against N-Ag in patients seropositive on admission.

**Fig. 3b.**
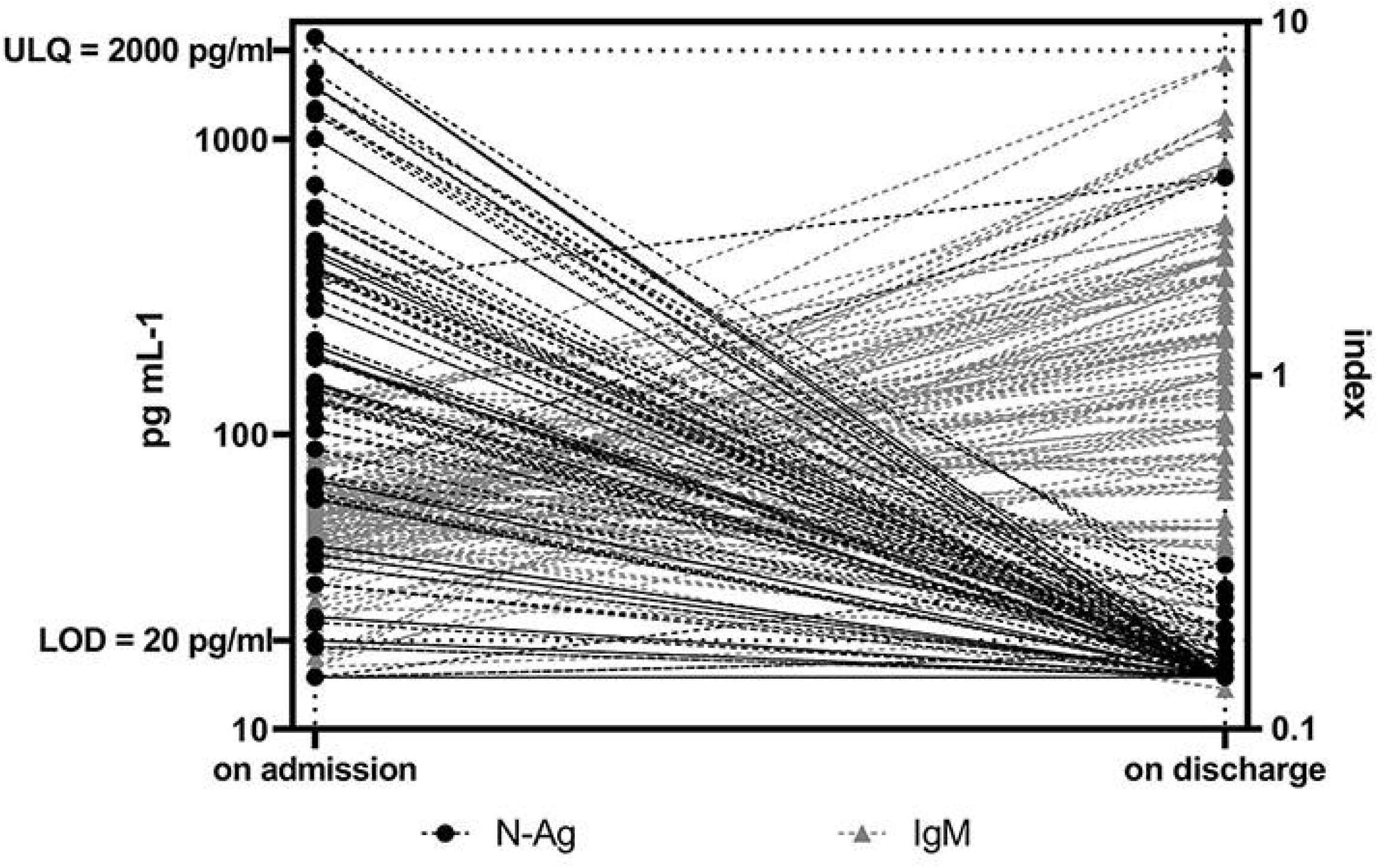
Kinetics of N-Ag and lgM antibodies against N-Ag in patients seropositive on admission.

Co-detection of N-Ag and anti-N-Ag antibodies in 92 (19%) of the clinical samples demonstrates that the epitope recognized by immunoassay pair used for the antigen determination is at least not fully overlapped (masked) by human antibody response.

For a subcohort of 20 patients, we had the opportunity to evaluate seroconversion within 5 days after the admission. The results shown in Figure 4 demonstrate that in the majority of cases the switch from Ag +Ab- to Ag-Ab+ status proceeded within 3-5 days after the emergency admission.

**Fig. 4.**
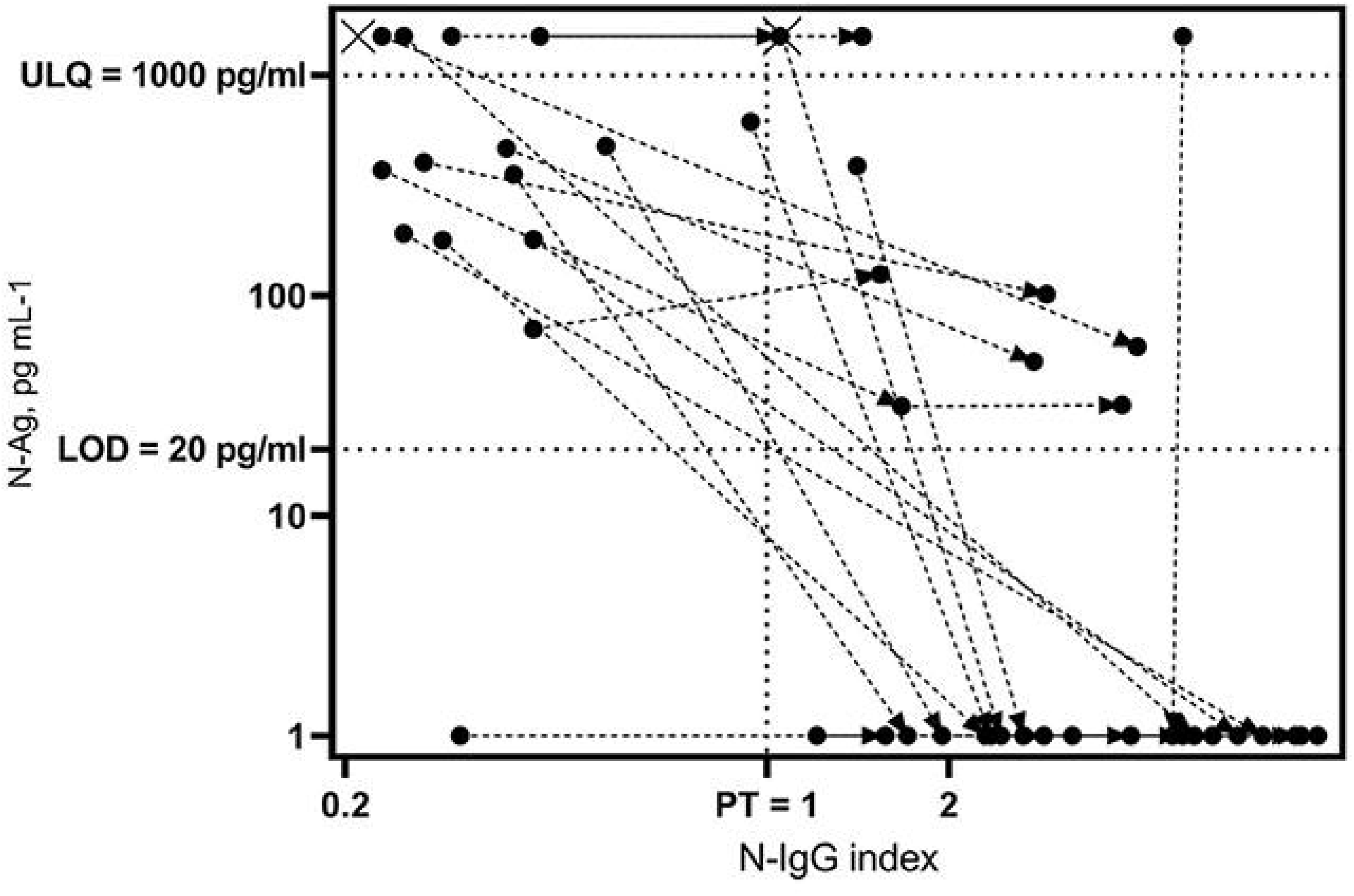
Individual seroconversion patterns within 1st week after admission.

The results indicate high relevance of combined serological tests for SARS-CoV-2 N-Ag and the antibodies to SARS-CoV-2 antigens as applied to the infectious pneumonia emergency patients at the admission, since:

- the sensitivity and specificity of the serological test for N-Ag is comparable to that of the RT-PCR analysis of nasopharyngeal swabs, and the use of serodiagnostics in combination with PCR tests for SARS-CoV-2 provides a significant increase in overall sensitivity;
- processing of blood samples is safer for the personnel due to the negligible presence of SARS-CoV-2 infectious particles in the blood, by contrast with nasopharyngeal swabs;
- serological tests are faster than PCR tests (when accounting for the full cycle of biomaterial processing and sample preparation), which accelerates the determination of SARS-CoV-2 status for a prompt decision on the management mode and isolation measures; in addition, blood serum analyses are easier to automate;
- collection of blood samples in this case is not an additional invasive procedure, but is performed as a part of the mandatory primary examination immediately upon the admission (as well as the final examination upon the discharge from the hospital).

According to the results of the study, hospitalization of the patients with SARS-CoV-2-associated pneumonia at the height of pandemic most frequently occurred before the onset of seroconversion (i.e. against the background of detectable serum N-Ag concentrations). In the majority of patients, the antigen prevailed upon admission and was “replaced” with respective antibodies by the time of discharge. Nevertheless, in some cases, at the time of hospitalization, the level of antibodies to N-Ag in the patient’s blood was already high, probably reflecting either delayed hospitalization or individual variation in the COVID-19 pathogenesis.

Overall, the obtained results indicate high feasibility of SARS-CoV-2 serodiagnostics in the emergency patients (along the lines of ‘rapid tests’ Capillus HIV-1/HIV-2®, Determine HIV-1/2®, etc. currently proposed as a gold standard of HIV diagnostics instead of the more expensive and time-consuming western blot analysis [9]). It should be noted that false-positive results of SARS-CoV-2 serodiagnostics usually reflect the presence of comorbidities associated with altered serological profiles (e.g. high levels of the IgG-binding rheumatoid factor in rheumatoid arthritis or the presence of HAMA antibodies), which underscores the significance of personalized comprehensive diagnostics for the patients with severe COVID-19.

## Data Availability

The authors confirm that the data supporting the findings of this study are available within the article and its supplementary materials.

## Author Contributions

YSL designed the study, performed the sampling, conducted molecular studies and drafted the manuscript, OVL and AGG performed the clinical examinations and sampling, conducted biochemical and molecular studies, AVP conducted molecular studies and drafted the manuscript, VVB and DVR designed the study and drafted the manuscript. All authors read and approved the final version of the manuscript.

## Ethics Approval and Consent to Participate

The study protocol was reviewed and approved by the Local Ethics Committee of the Pirogov Russian State Medical University (meeting No.194 from March 16 2020, Protocol No.2020/07); the study was conducted in accordance with the Declaration of Helsinki.

